# The impact of age, comorbidity, and current medication use on plasma p-tau217 in adolescents

**DOI:** 10.64898/2026.03.30.26349647

**Authors:** Stephani L. Stancil, Mariah E. Brewe, Hana Mayfield, Jill Morris

## Abstract

**Background:** Adolescence is a critical period of neurodevelopment with the emergence of chronic medical conditions and increasing exposure to long-term medications. P-tau217 is a sensitive blood-based biomarker of neuropathology in older adults, yet its developmental behavior and susceptibility to common clinical factors in youth are unclear. Here we tested whether p-tau217 varies with age, comorbidity, or medication use during adolescence; and whether collection method (venous vs Tasso+ capillary) yields comparable concentrations.

**Methods:** In an adolescent cohort, plasma p-tau217 was measured by Simoa HD-X. Paired venous and Tasso+ capillary samples were also analyzed from adult volunteers for methodological comparison

**Results:** In adolescents (n=41; mean age 16±2.6 years), p-tau217 did not correlate with age or BMI z-score and did not differ by psychiatric, cardiometabolic, or gastrointestinal comorbidity, nor by corresponding medication use. In contrast, p-tau217 concentrations were >10-fold higher in Tasso+ capillary plasma than venous plasma, a discordance replicated in paired adult samples.

**Conclusion:** Plasma p-tau217 appears physiologically stable across common clinical variables in adolescence, but highly sensitive to biospecimen collection method. Venous and Tasso+ capillary plasma should not be directly compared or pooled until methodological differences are resolved. These data provide a developmental baseline and critical methodological caution for pediatric neuroscience and decentralized biomarker studies.

## INTRODUCTION

Plasma phosphorylated tau at threonine 217 (p-tau217) has emerged as one of the most sensitive and specific blood-based indicators of Alzheimer’s Disease (AD)–related neuropathology in adults, where elevated concentrations reliably track amyloid and tau PET burden, cortical atrophy, and cerebrospinal fluid (CSF) markers of neurodegeneration^1-4^, even before clinical cognitive impairment is clinically apparent^5^. While often treated as AD-specific, tau phosphorylation reflects broader neuronal processes - altered microtubule binding affinity and axonal transport - that could, in theory, vary across development and indicate neuronal vulnerability^6^. Defining physiological and clinical sources of variation outside of neurodegeneration in youth is therefore essential.

Adult studies suggest that p-tau217 is relatively robust to many common medical conditions and medication exposures. Evidence suggests that renal function (e.g., creatinine, eGFR), hemoglobin levels, and indices of metabolic health show only small associations with p-tau217^7-9^ though some studies suggest renal function may affect levels^10,11^. Compared with other blood-based neurological biomarkers, p-tau217 shows minimal susceptibility to confounding by BMI, lipids, thyroid function, glucose regulation, cardiovascular comorbidities or their treatments^8,12^. Although tau-related processes have been implicated in certain psychiatric conditions, including depression^13,14^, nearly all available evidence comes from middle-aged or older adults.

Limited developmental data indicate that newborns exhibit markedly elevated p-tau217 in venous cord blood that declines to low concentrations within the first year, whereas sparse data in adolescents (n=12) show values similar to cognitively normal older adults—raising the possibility that p-tau217 participates in neurodevelopmental biology as well as neurodegeneration, underscoring the need to define its behavior during adolescence^15^.

Adolescence represents a critical developmental period characterized by dynamic neurobiological maturation as well as the emergence of chronic health conditions and initiation of long-term medications. Establishing whether age, comorbidities, or commonly used treatments influence p-tau217 during this stage is essential both for interpreting biomarker findings in pediatric research and for laying groundwork for potential future clinical applications in at-risk youth. Here, we address two questions: (1) does plasma p-tau217 vary with age, common comorbidities, or current medication use during adolescence; and (2) are concentrations comparable across venous and Tasso+ capillary plasma? By delineating physiological stability and identifying methodological pitfalls, we aim to provide a developmental baseline and practical guidance for decentralized biomarker research.

## METHODS

### Participants

Cross-sectional cohort of adolescents aged 10-21 years old receiving outpatient care in the

Division of Adolescent and Young Adult Medicine at a pediatric academic medical center and enrolled in a biorepository. Demographics, medical history and current medication use were obtained by self-report and review of electronic medical records, then grouped into common categories (see **Table 1**). Blood was obtained by venipuncture and collected in lavender EDTA tubes. Plasma was separated by centrifugation (1000g x 10 minutes) and stored at -80 within 1 hour of collection.

**Table 1.**
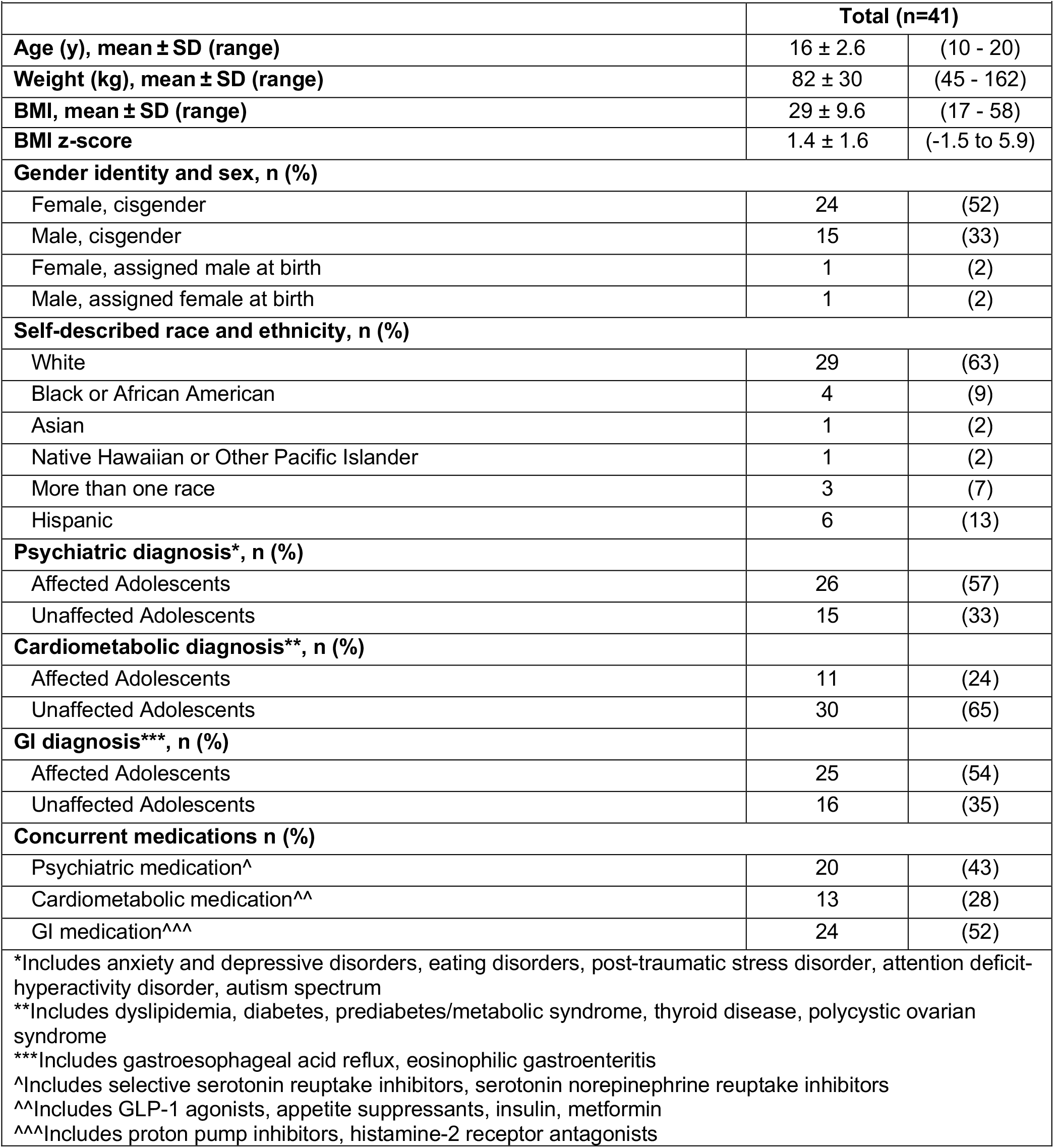
Participant Characteristics.

### Ethics approval

This research was reviewed and approved by the Institutional Review Board at Children’s Mercy Kansas City and informed permission/assent/consent was obtained from all participants.

### Paired samples

Paired venous and Tasso+ capillary plasma samples were obtained from six adult volunteers as part of assay and collection optimization procedures. Samples were de-identified and analyzed as non-human subjects research.

### Plasma p-tau217 assay

Immediately prior to analysis, plasma samples were thawed at room temperature for 75 minutes, then vortexted ∼8 seconds prior to centrifugation at 10,000 x g for 5 minutes. 110uL of plasma was pipetted into each sample well and analyzed on a Quanterix Simoa HD-X using the ALZPath pTau217 Advantage Plus kit according to manufacturer instructions. Plasma p-tau217 measurements in Cohort 1 were done in duplicate and reported as mean value. The mean CV was <5% in all runs. Signal variations within and between analytical runs were assessed using two internal quality control plasma samples at the beginning and the end of each run. Plasma samples had undergone a maximum of 2 freeze-thaw cycles prior to analysis. All sample values were above the functional lower limit of quantification for the assay.

### Statistical analysis

All statistical analyses were conducted using R (Version 2025.05.0, Posit, Inc). Normality was assessed using Q-Q plots and the Shapiro-Wilk test. Since p-tau217 data were found to be normally distributed, descriptive statistics were reported as mean ± standard deviation and correlations were evaluated using Pearson’s r. Group differences were analyzed using Welch’s t-test. Effect sizes were assessed with Hedges’ g. Two-tailed p values <0.01 were considered significant (conservative adjustment for multiple comparisons). To examine the association between plasma p-tau217 and individual factors, linear regression fit three models as follows: (1) model 1 adjusted by age, BMI z-score, and sex; (2) model 2 adjusted by the variables in model 1 plus presence or absence of psychiatric, cardiometabolic, and GI comorbidity; and (3) model 3 adjusted by the variables in model 2 plus current medication use grouped by medication class (see **Table 1**). Collinearity among variables was assessed with variable inflation factor (VIF). Model selection was guided by Akaike Information Criterion.

## RESULTS

**Table 1** lists participant demographics including comorbidities and concurrent medications grouped by category as well as individual diagnoses and medications within categories.

### Plasma p-tau217 by age, comorbid conditions, and current medication use in adolescents. Figure 1

shows lack of correlation between p-tau217 and age during the second decade of life (r = -0.10 [95%CI -0.40 to 0.21], p=0.5266) or between p-tau217 and BMI z-score across a wide range from underweight to obese (r = -0.10 [95%CI -0.40 to 0.23], p=0.5638).

**Figure 1.**
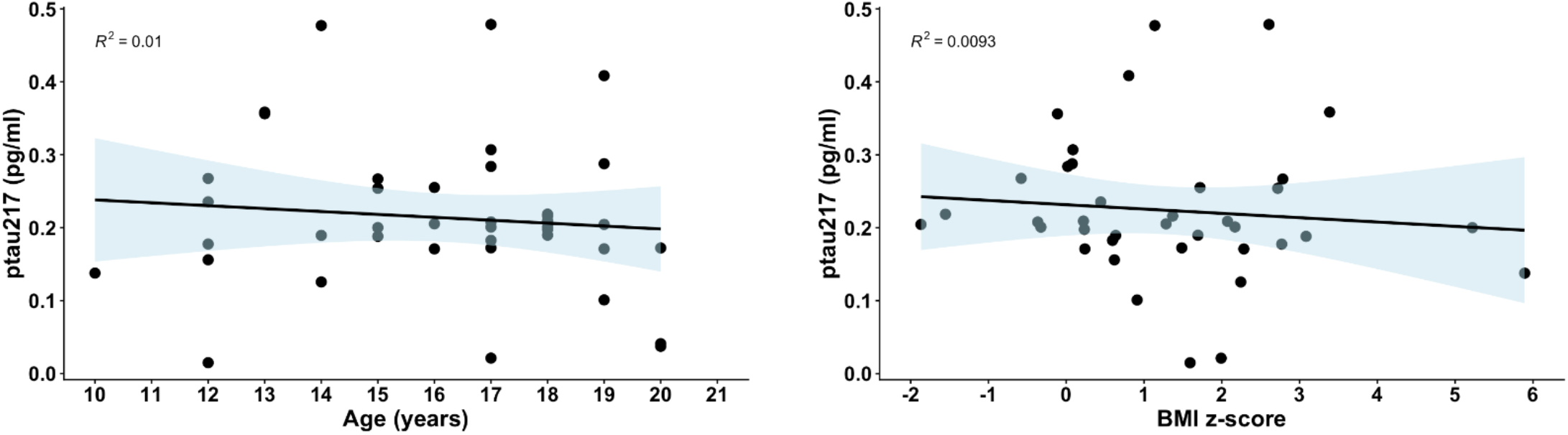
Plasma p-tau217 as a function of age and BMI z-score. In adolescents, p-tau217 values were not associated with age (r = -0.10 [95%CI -0.40 to 0.21], p=0.5266) or BMI z-score (r = -0.10 [95%CI -0.40 to 0.23], p=0.5638).

Plasma p-tau217 did not differ by the presence or absence of psychiatric, cardiometabolic, or gastrointestinal comorbidities (**Figure 2**). Mean±SD plasma p-tau217 by comorbidity were as follows: psychiatric diagnosis: affected (n=26) 0.23±0.10, unaffected (n=15) 0.19±0.11, p=0.278, Hedges’ g= -0.36 [-0.99 to 0.27]; cardiometabolic diagnosis: affected (n=11) 0.16±0.06, unaffected (n=30) 0.23±0.11, p=0.017, Hedges’ g= 0.67 [-0.03 to 1.36]; GI diagnosis: affected (n=25) 0.23±0.11, unaffected (n=16) 0.19±0.09, p=0.178, Hedges’ g= - 0.42 [-1.04 to 0.21]. Similarly, plasma p-tau217 did not differ significantly among adolescents taking antidepressants, cardiometabolic medications or GI medications compared with those not taking these agents (data not shown).

**Figure 2.**
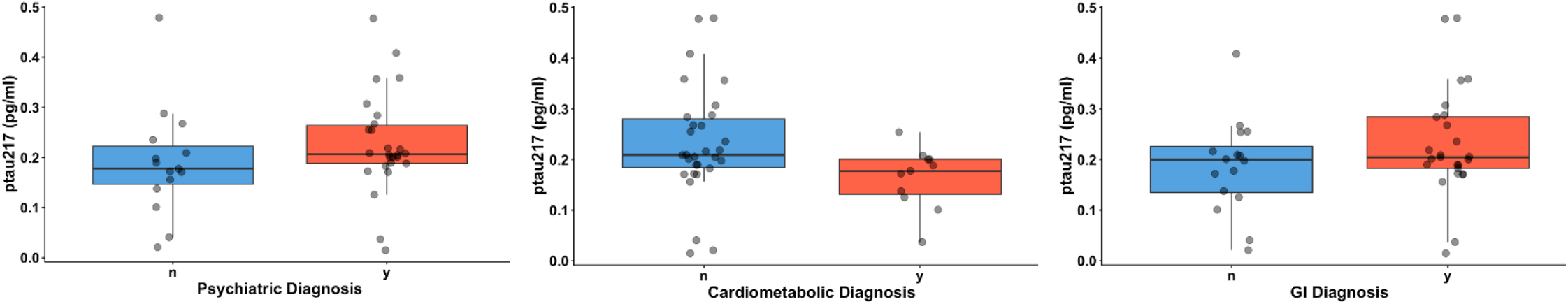
Individual p-tau217 values in adolescents with medical comorbidities. Boxplots represent group means of those unaffected (n) and affected (y) with psychiatric, cardiometabolic, and GI comorbidities, respectively. All comparisons non-significant (Welch’s t-test).

### Impact of individual factors on plasma p-tau217 in adolescents

Linear regression models 1-3 (**Figure 3**) did not find any significant influence of age, sex, BMI z-score, comorbidity, or current medication use on plasma p-tau217 in adolescents aged 10-21 years (adjusted R^2^ = -0.06 to -0.04; p = 0.817 to 0.541; AIC -60.2 to -56.2). Covariates did not show evidence of collinearity (variance inflation factors <2.8) with the exception of GI diagnosis and GI medication in model 3 (VIFs 13). Removing GI diagnosis or GI medication use from model 3 resolved the collinearity with resulting VIFs <2.4 for remaining variables. Model 3 remained non-significant, and AICs were similar whether GI diagnosis or GI medication use was retained (AIC -57.8 and -58.1, respectively).

**Figure 3.**
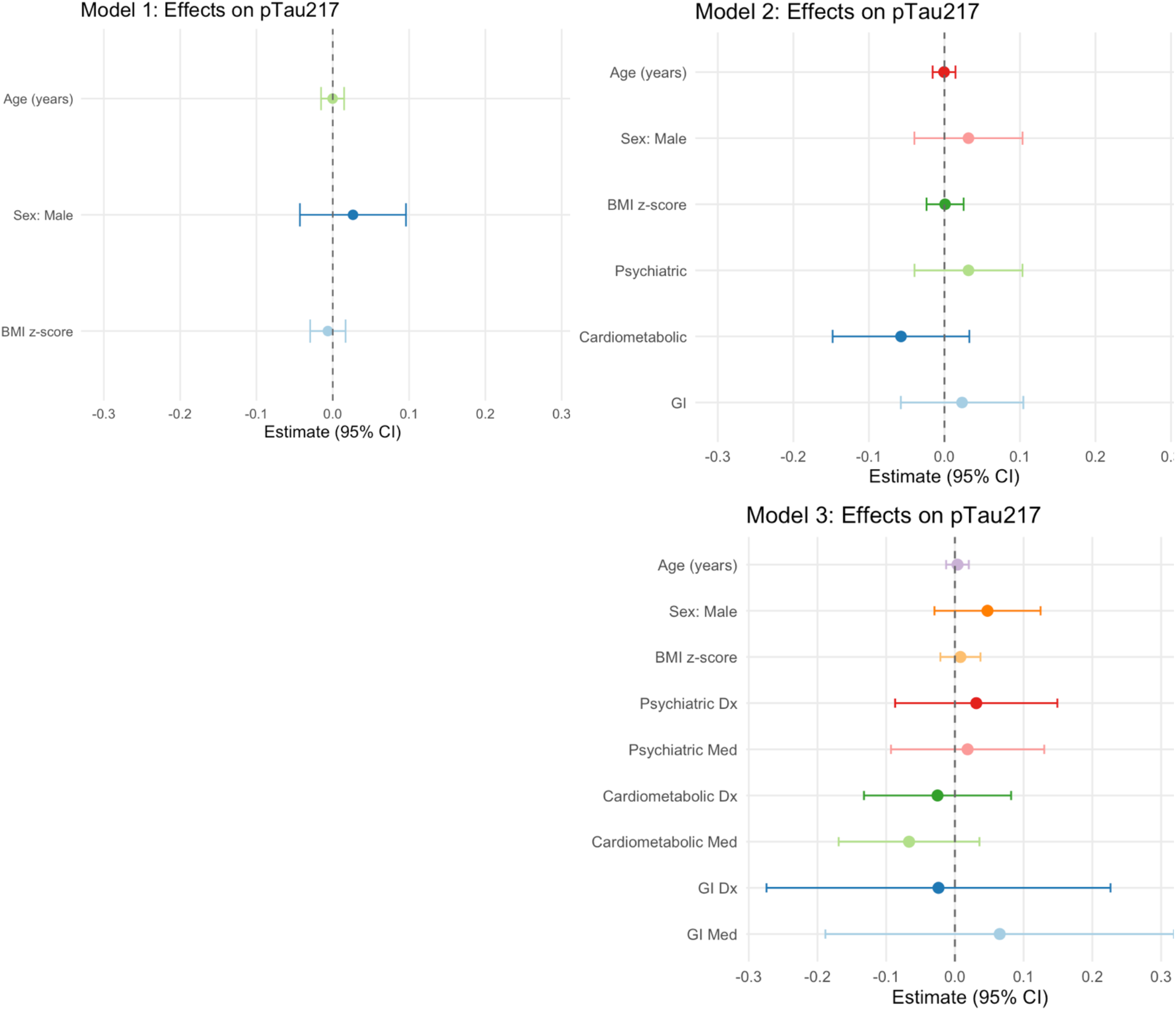
Age, sex, BMI z-score, common comorbidities and current medication use does not influence p-tau217 in adolescents. Forest plots represent beta estimates from linear regression models evaluating the impact of base covariates, medical history, and medication use on plasma p-tau217 in adolescents aged 10-21 years old. Model 1: predictor + age + sex + BMI z-score. Model 2: predictor + age + sex + BMI z-score + psychiatric diagnosis + cardiometabolic diagnosis + GI diagnosis. Model 3: predictor + age + sex + BMI z-score + psychiatric diagnosis + psychiatric medication + cardiometabolic diagnosis + cardiometabolic medication + GI diagnosis + GI medication. Models 2 and 3 evaluated the presence or absence of comorbidity and current medication use.

### Methodologic finding of discordance between venous and capillary collected plasma p-tau217

Five participants were excluded from the primary analysis due to substantially elevated p-tau217 values in plasma collected by the Tasso+ capillary collection device. Both Tasso+ and venipuncture samples were collected into lavender EDTA tubes and sample processing was consistent; however, mean plasma p-tau217 values were >10-fold higher in Tasso+ capillary samples (Tasso+ capillary: 4.51±1.17, n=5; venous: 0.21±0.10, n=41). This observation was replicated in paired venous and Tasso+ capillary samples from 6 healthy adults. Plasma p-tau217 was 3.73±1.58 and 0.40±0.15 from Tasso+ capillary and venous samples, respectively, with a non-significant negative correlation (r = -0.35 [95%CI -0.90 to 0.65], p=0.503; **Figure 4**).

**Figure 4.**
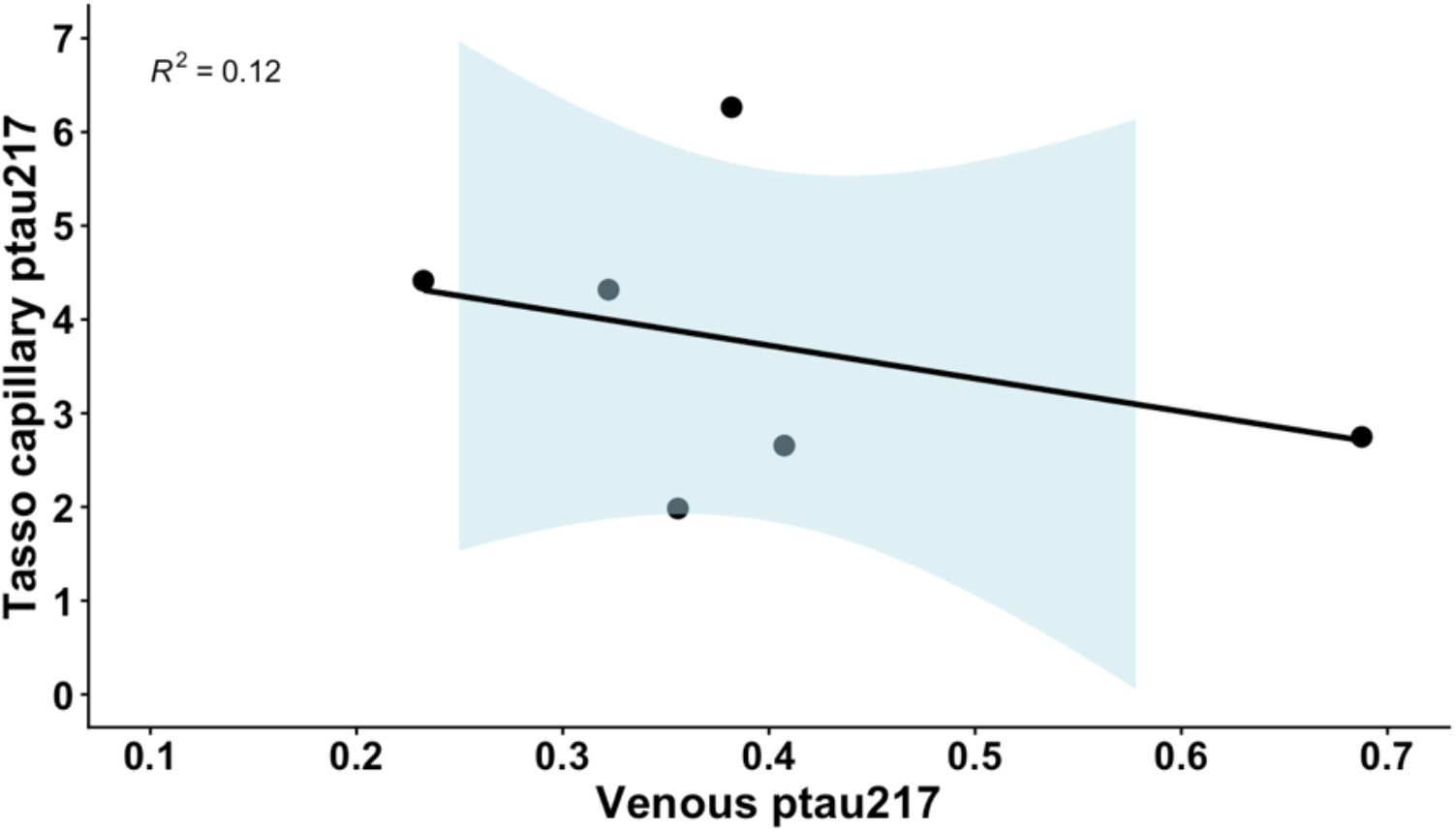
Relationship between p-tau217 measured in Tasso+ capillary and venous samples. Methodological comparison of matched samples from healthy donors. P-tau217 reported as pg/ml.

## DISCUSSION

This study characterizes plasma p-tau217 concentrations in adolescents and establishes a baseline understanding of how age, common comorbidities, and current medications relate to this biomarker during a key developmental period. In contrast to the physiologic stability observed across clinical variables, plasma p-tau217 concentrations differed strikingly by blood collection method.

Overall, venous plasma p-tau217 concentrations in this adolescent cohort were low and largely within the “normal” range established in older adults (>65 years). Only 7% (n=3) of participants exceeded the conservative threshold of 0.40 pg/ml, and none exceeded the higher-risk threshold of 0.63 pg/ml that has shown high negative and positive concordance with cerebral amyloid status^4^. Although small pediatric studies have reported elevations in total tau or alternative phosphorylated tau species (e.g., p-tau181) in central nervous system disease or ADHD^16-19^, whether such conditions influence p-tau217—arguably the most analytically robust tau biomarker—had not been established prior to this work.

P-tau217 is theoretically promising as a proxy of healthy neuronal signaling; however, its utility in pediatric patients has not been established. The specific context of use needs to be carefully considered particularly since the robust performance of p-tau217 is centered on prediction of brain amyloid positivity (by PET) associated with cognitive impairment in older adults (>65 years). One might imagine future potential uses in pediatrics such as early identification of a high-risk phenotype for cognitive impairment (and opportunity for intervention); however, this exploration in children and adolescents, which would involve PET use, carries ethical and safety considerations not the least of which is PET-related ionizing radiation exposure. Recent research suggests lower p-tau217 thresholds hold optimal sensitivity and specificity for cognitively normal adults less than 65 years, with 0.35 pg/ml noted to have 85% sensitivity and 95.7% specificity for PET amyloid positivity^20^. Notably, 12% (n=5) of adolescents in our cohort exceeded this threshold. Whether such values reflect amyloid positivity in adolescents or whether amyloid positivity is an appropriate marker of neural disturbance in younger populations remains unclear. More immediate and developmentally relevant applications for plasma p-tau217 may lie in research focused on elucidating neural mechanisms of psychiatric disease rather than direct disease prediction.

Sample collection and processing are critical for interpreting p-tau217. Validated protocols support the use of EDTA tubes and analysis within three freeze–thaw cycles^4,21^, with strong reliability across most assay platforms^22^. Our venous samples adhered to these best practices and were analyzed within two freeze–thaw cycles. In contrast, we observed that capillary-derived p-tau217 concentrations were substantially higher than paired venous values, a difference not previously appreciated. Prior work has examined agreement between capillary dried blood spot and venous samples^23^ and between Tasso+ capillary samples and CSF^24^, but no study has directly compared p-tau217 concentrations in venous versus Tasso+-collected capillary plasma as done here. While Tasso+ has demonstrated concordance with venous sampling for other analytes^25^, the discrepance in p-tau217 has immediate relevance for decentralized trial designs and emerging direct-to-consumer biomarker applications (e.g., Neurogen).

This study has limitations. The cross-sectional design was well suited to establishing baseline distributions in adolescents and evaluating associations with age, common comorbidities, and medication use. Our study was not designed to address within-person variability or responses to acute CNS perturbations (e.g., head injury, infection) and we did not analyze kidney function, a clinical variable that may affect biomarker outcomes. The comorbidities were grouped into the most common diagnoses to enable analyses (psychiatric, cardiometabolic, and GI diagnoses). Expectedly, most common medication classes were aligned with the most common diagnostic groups. Detecting differences attributable to an individual medication or diagnosis is outside the scope of this study. Whether adolescent p-tau217 is sensitive to acute CNS injury, or how genetic and familial risk factors for neurodegeneration (e.g., APOE ε4, Alzheimer’s family history) influence p-tau217 during youth, remains unknown.

In conclusion, these findings indicate that plasma p-tau217 is physiologically stable during adolescence but highly sensitive to sample collection methodology. Venous and Tasso+ capillary plasma should not be directly compared or pooled until methodological differences are fully understood. As p-tau217 moves rapidly into decentralized trials and consumer-facing testing, rigorous attention to biospecimen type will be essential to avoid misinterpretation.

## Data Availability Statement

Data are available upon reasonable request to the principal investigator and following relevant regulatory approval(s).

